# Risk and outcomes of healthcare-associated infections in three hospitals in Bobo Dioulasso, 2022 (Burkina Faso): a longitudinal study

**DOI:** 10.1101/2024.07.04.24309979

**Authors:** Arsène Hema, Satouro Arsène Some, Odilon Kaboré, Soufiane Sanou, Armel Poda, Ziemlé Clément Meda, Abdoul Salam Ouedraogo, Léon. Savadogo

**Affiliations:** Quality Department of Sourô Sanou Teaching Hospital, Bobo-Dioulasso, Burkina Faso; Centre Muraz, National Public Health Institute, Bobo-Dioulasso, Burkina Faso; Bacteriology and virology laboratory, Bobo-Dioulasso, Burkina Faso; Infectious Diseases Department of Sourô Sanou Teaching Hospital, Bobo-Dioulasso, Burkina Faso; Information, Research and Epidemiology Department of Sourô Sanou Teaching Hospital, Bobo-Dioulasso, Burkina; National Institute of Health Sciences, Nazi Boni University, Bobo-Dioulasso, Burkina Faso

**Keywords:** longitudinal study, Burkina Faso, multicentre, healthcare-associated infection, outcomes

## Abstract

**Background:** Healthcare-associated infections (HAIs) are one of the most common adverse events in healthcare and represent a major public health problem. However, 40% to 70% of HAIs are considered to be preventable. The present study was conducted to analyze the incidence, etiological factors, and outcomes of HAIs through active surveillance in three hospitals in the city of Bobo Dioulasso.

**Methods:** A prospective, longitudinal, multicenter study was conducted from May 1^th^ to November 30^rd^, 2022, in two district hospitals (DO and Dafra) and the Sourô Sanou Teaching Hospital (CHUSS). Consenting patients hospitalized for reasons other than infection, cancer, immunosuppression in the postoperative care ward of DO or of Dafra district hospitals, intensive care unit (ICU)/CHUSS, neonatal ward/CHUSS, and gynecology and obstetrics postoperative care ward/CHUSS during a 2-month inclusion period in district hospitals and 4 months for CHUSS wards. For this study, we used the operational definitions of the French Technical Committee for Nosocomial Infections and Healthcare-associated Infections, with slight modifications. Logistic regression was used to analyze predictors of HAIs.

**Results:** Of the 664 patients enrolled, 166 experienced an HAI, with a cumulative incidence rate of 25% (CI: 21.7%-28.3%) or an incidence density rate of 36.7 per 1000 patient-days (CI: 31.7-42.9). Surgical site infections (SSI) (44%), followed by neonatal infections (42%) were the most common HAIs. Enterobacteriaceae represented 60% of the bacteria identified in HAIs, and 38.9% of them were extended spectrum β-lactamase (EBLSE) producers. Factors associated with HAIs were admission in the neonatal ward (aOR=7.4; CI:1.3-42.7), ICU (aOR=3.7; CI:1.4-9.5), hospital stay longer than 2 days (aOR=2.1; CI:1.2-3.4), or male sex (aOR=1.8; CI:1.1-3.1). In addition, HAIs were associated with longer follow-up, hospitalization, and mortality (18.1%; 95% CI:12.1 - 24.4). Deaths were only recorded in the ICU and neonatal ward, with case fatality rates of 45.4% (95% CI: 27.5 - 63.4) and 21.4% (95% CI: 11.6 - 31.3), respectively, p=0.019.

**Conclusions:** The incidence of HAIs was relatively high in the three hospitals in Bobo Dioulasso. A national strategy to reduce HAIs should be implemented to achieve better control of HAIs.

## Introduction

Healthcare-associated infections (HAIs) are one of the most common adverse events in healthcare and a major public health problem in both developed and resource-limited countries [1]. HAIs are associated with increased healthcare costs, prolonged hospital stays, functional disability, and patient death [1,2]. Most HAIs are caused by bacteria, viruses or microscopic fungi that become multi-resistant due to inappropriate use of antimicrobials. Thus, multi-resistant microorganisms cause infections with limited therapeutic options, leading to longer hospital stays and higher mortality rates compared to HAIs caused by susceptible bacteria of the same species [2–5]. Therefore, the fight against HAIs is inextricably linked to the fight against antimicrobial resistance [1,6]. Many factors promote HAIs, including healthcare systems and procedures, human behavior influenced by education, socioeconomic constraints, and often societal norms and beliefs. However, 40% to 70% of HAIs are considered preventable through adherence to hospital hygiene measures [7,8].

According to the World Health Organization (WHO), HAIs affect 1.4 million people worldwide at any given time, with 3.5% to 12% in developed countries (9% in the United Kingdom, 4.4% in France, and 6.9% in Belgium) compared to 25% in low- and middle-income countries [1,2]. In Africa, hospital-wide prevalence of HAIs varied from 6.7% to 18.7%, with a high prevalence that can exceed 50% in ICUs [9–13]. Most of these studies are prevalence studies, which have the advantage of being easy and quick to conduct, but the disadvantage of low precision and lack of information on the outcomes of HAIs [14].

In Burkina Faso, data on the incidence of HAIs are scarce due to the lack of an HAI surveillance system [15, 16]. In addition, hospital overcrowding, limited resources, structural and logistical challenges, combined with inadequate training of professionals in hospital hygiene, antimicrobial resistance and their inadequate monitoring, suggest a high risk of HAIs. The present study aims to analyze the extent, etiologic factors and outcomes of associated infections through active surveillance in three hospitals the city of Bobo Dioulasso.

## Methods

### Study design and location

This prospective, longitudinal, multicenter study was conducted from May 1^th^ to November 30^th^, 2022, in 3 health care facilities in Bobo-Dioulasso, the second largest city in Burkina Faso. The facilities included two (02) district hospitals (DO and Dafra) and one (01) teaching hospital (Sourô Sanou Teaching Hospital (CHUSS) in Bobo-Dioulasso. In the district hospitals, the study was conducted in the postoperative care wards. In the Sourô Sanou teaching hospital, the study was conducted in the intensive care unit, the gynecology and obstetrics postoperative care ward and the neonatal ward.

#### Postoperative care ward of Dafra district hospital

The post-operative ward of Dafra District Hospital received 888 post-operative patients in 2022. This ward consists of two (02) inpatient rooms with a total capacity of 16 beds. During the survey, the service was staffed by 52 nurses, two (02) hospital hygiene technicians, and two permanent physicians.

#### Postoperative care ward of Do district hospital

The postoperative ward of Do District Hospital received 1706 postoperative patients in 2022. This ward consists of six inpatient rooms, with a total capacity of 24 beds. At the time of the survey, the service was staffed by 59 nurses, 1 hospital hygiene technician, five (05) permanent physicians, and approximately five (05) medical trainees.

#### Intensive Care Units of Sourô Sanou Teaching Hospital (ICU)

It is the reference center for intensive care of four health regions out of 13 of Burkina Faso. In 2022, 525 patients were admitted to the intensive care unit of Sourô Sanou Teaching Hospital. This unit has 14 inpatient beds. The service is staffed by five (05) Physicians Anaesthetist resuscitators, approximately 10 physicians’ residents, 23 nurses and three (03) hospital hygiene technicians

#### Neonatal ward of Sourô Sanou Teaching Hospital

It was the only neonatal ward in western Burkina Faso and a reference center for neonatal care of four health regions. In 2022, the neonatal ward of Sourô Sanou Teaching Hospital received 2,765 new-borns. It has a hospital room that includes 6 incubators and 34 cradles. The staff consists of 2 doctors specializing in pediatrics, 2 general practitioners, approximately 10 resident physicians, 28 nurses and 3 hospital hygiene technicians.

#### Gynecology and Obstetrics postoperative care ward of Sourô Sanou Teaching Hospital

In 2022, the gynecology and obstetrics postoperative care ward of Sourô Sanou Teaching Hospital received 1,946 women, including 1,433 cesareans. This unit has 30 inpatient beds. The staff consisted of 16 physicians, four (04) nurses and 21 midwives.

### Study population

The population of this study consisted of the cohort of all patients admitted to the neonatal ward, ICU or the gynecology and obstetrics postoperative care ward of Sourô Sanou Teaching Hospital, as well as the postoperative care ward of Do District Hospital and of Dafra District Hospital during the period of active HAI surveillance.

### Inclusion criteria

Were included in this study, patients meeting the following conditions:

– Admitted to the postoperative care ward of Do or Dafra District Hospital, from May 1^th^ to June 30^rd^, 2022 or

– Admitted to ICU or neonatal ward of Sourô Sanou Teaching Hospital, from May 14^th^ to August 14^th^, 2022, or

– Admitted to the gynecology and obstetrics postoperative care ward of Sourô Sanou Teaching Hospital, from August 1^th^ to November 30^rd^, 2022

– and have given their consent

### Non-inclusion criteria

Patients were not included in this study if they met any of the following criteria:

– Patients who have been hospitalized for less than 48 hours,

– Had a primary diagnosis of sepsis, cancer, infection, or immunocompromised state.

### Surveillance and data collection

Based on data from a previous prevalence study [16], we targeted 4 infections to monitor for this study: urinary infections, systemic infections, surgical site infections (SSI) and neonatal infections. A team composed of an epidemiologist, two clinical doctors and three nursing managers was responsible for active data collection in collaboration with the care unit supervisor. Data were collected from patient records, bandage records, hospitalization records and patient interviews. Case identification was active. For this purpose, the data collection team examined hospitalization registers three times a week to identify eligible patients. Information collected from these sources was supplemented by information from patients and nursing staff and recorded on individual data collection sheets which included, among others: sociodemographic data (age, sex, origin, profession), factors risk related to care, initial or curative antibiotic therapy, HAI type, pathogen in question, development. Surgical patients included were followed until the surgical wound healed, even after discharge from the hospital. The condition of the surgical wounds was monitored during outpatient bandage visits. Non-operated patients were followed until they left the hospital. In all cases, patient follow-up did not exceed 30 days. In the event of HAI, biological samples when possible were taken when possible, depending on the type of suspected infection (blood, pus, urine) and a bacteriological analysis were performed in the CHUSS laboratory.

### Definition of outcome variables

The primary outcome variable was occurrence of infection after 48 h of hospitalization in a patient with no symptomatic or incubating infection at the time hospital admission. For this study, we used the operational definitions of the French Technical Committee for Nosocomial Infections and Healthcare-Related Infections (CTINILS) [17], with slight modification. This committee largely adopts the definitions of the Center of Disease Control and prevention (CDC).

### Variables Outcome variables

Outcome variables were HAI and its subsequent outcome. HAI was defined by the presence of at least one of the infections targeted by surveillance during this study (SSI, urinary tract infection, systemic infection, neonatal infection). For the analysis, we coded: 0 for no HAI and 1 for presence of HAI. HAIs outcomes were categorized by recoveries, deaths, discharges against medical advice and lost to follow-up.

### Predictor variables

– Age: categorized into four groups: 0-14 years, 15-35 years, 36-50 years, and >50 years.

– Gender: female and male

– Residence: the patient’s place of residence, categorized as urban or rural

– Exposure: Surgery, urinary catheter, peripheral venous catheter, length of stay in hospital before inclusion in the study (in days)

– Ward/Unit: these are the wards/units from which patients were recruited. There were three Wards/Units of Sourô Sanou teaching Hospital (ICU/CHUSS, Neonatology/CHUSS Gyn & Ob/CHUSS), one ward of the district hospital of Dafra (Surgical /Dafra) and one of the district hospital of DO (Surgical /Do).

– Follow-up duration: period between inclusion in the study and HAIs outcome occurrence. The duration of follow-up does not exceed 30 days

– Length of stay before inclusion the time spent in hospitalization and inclusion in the study in days

### Statistical analysis

Data collected on survey forms were entered into Epidata^®^ software and analyzed using Stata® software version 13. Patient characteristics, HAI exposure, and distribution of patients by ward were described. Incidence rates with their 95% confidence intervals (CI), type, antimicrobial use and HAI outcomes were described by patient characteristics, HAI exposure and ward/unit. Qualitative variables were described by their percentages and the quantitative variables by their mean (me) and their standard deviation (SD). Proportions were compared using Pearson’s chi2 test or Fisher’s exact test when appropriate. Means were compared using Student’s t-test. Logistic regression (univariate and multivariate) was used to identify the risk factor for HAI. Patient characteristics, HAI exposures and ward/unit that were significantly associated with HAI at the 20% univariate threshold were retained in the multivariate models. The significance threshold for all tests was set at a value of p < 5%.

### Ethical aspects

This study was conducted as part of the “Surveillance and prevention strategies for healthcare associated infections in Bobo-Dioulasso, Burkina Faso” study approved by the National Ethics Committee of Burkina Faso. Signed informed consent was obtained from the participants and an information sheet was given to each participant. We obtained informed consent, written and signed from parents or guardians for minors. The data collected were anonymized to ensure confidentiality.

## Results

### 3.1. Description of the study population

A total of 664 patients were included with 471 (70.9%) at the Sourô Sanou Teaching Hospital, 85 (12.8%) at the CMA of Dafra and 108 (16.3%) at the CMA of DO. The study population was mainly female (80.1%) with a mean age of the study population of 21.6 years (SD: 14.3). Neonates (less than 1 month) represented 25% of our study population. Our patients mainly lived in urban areas (88.7%). Regarding exposure, 69.9% of them had undergone surgery, 47.1% had a urinary catheter and 50.0% had a venous catheter (Table 1), and the mean length of hospital stay before inclusion was 2.2 days (SD: 0.4). The majority (34.9%) of the included patients were admitted to the gynecology and obstetrics postoperative ward of the CHUSS.

**Table 1:**
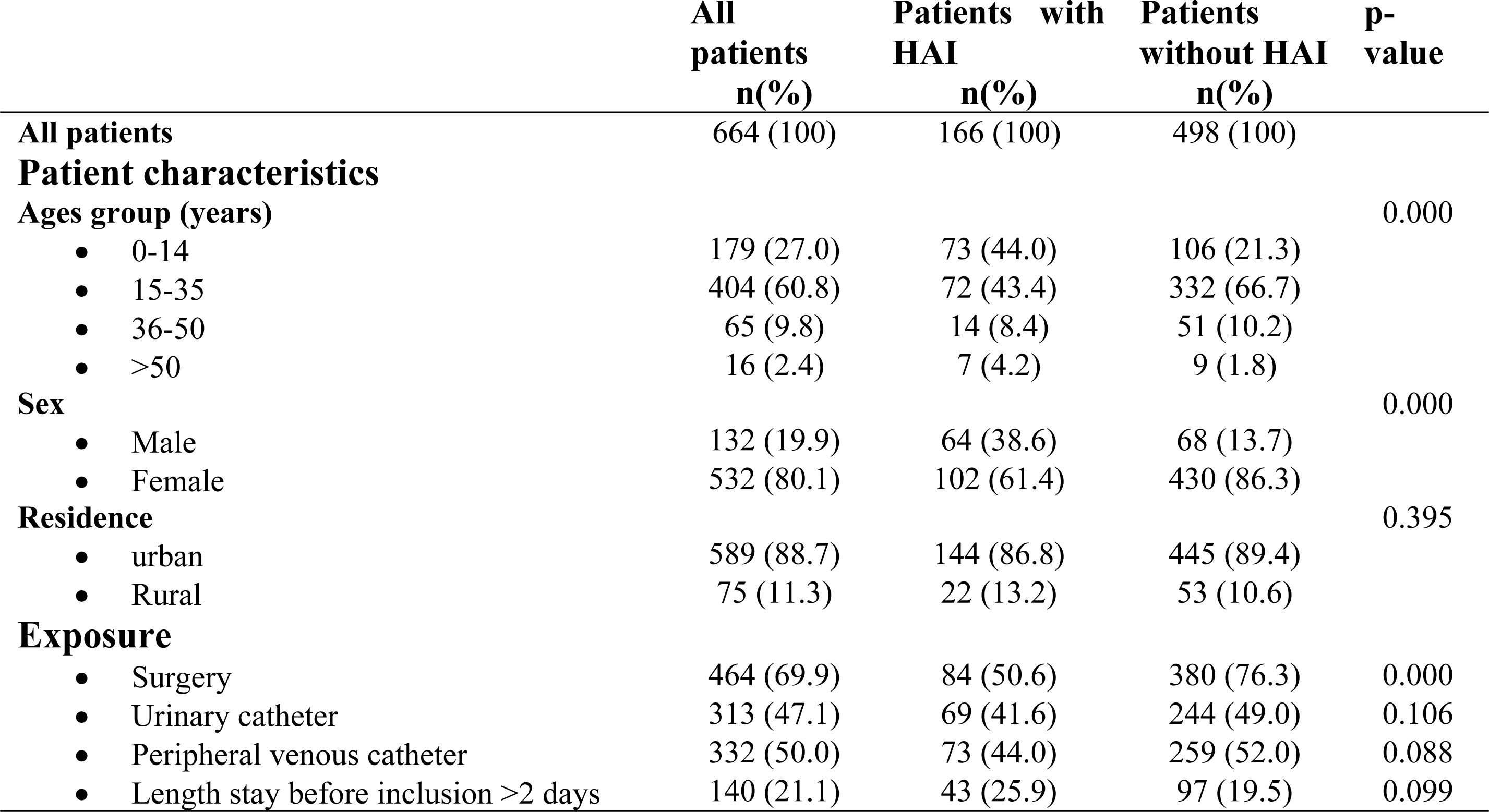
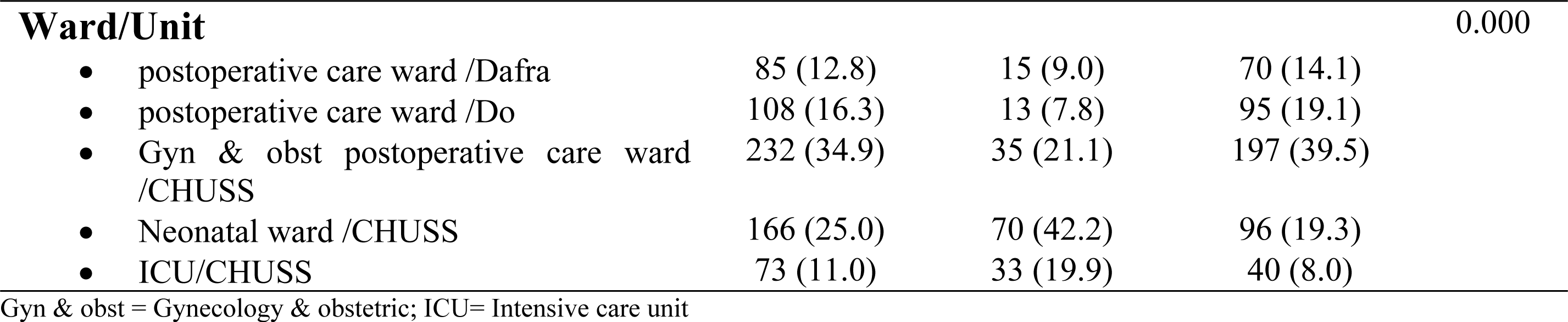
Characteristics of patients, multicenter longitudinal survey, Bobo-Dioulasso 2022.

### 3.2. Healthcare-associated incidence

Of the 664 patients followed, 166 had at least one HAI, resulting in a cumulative incidence rate of 25% (CI: 21.7%-28.3%) or an incidence density rate of 36.7 per 1000 patient-days (CI: 31.7-4.3). The mean time of the occurrence of these infections was 5 days (SD: 3.5). The highest incidences of HAI were observed in ICU (45.2%; CI: 33.5-56.9%) and the neonatal ward (42.2%; CI: 34.6-49.7%) (Table 2).

**Table 2:**
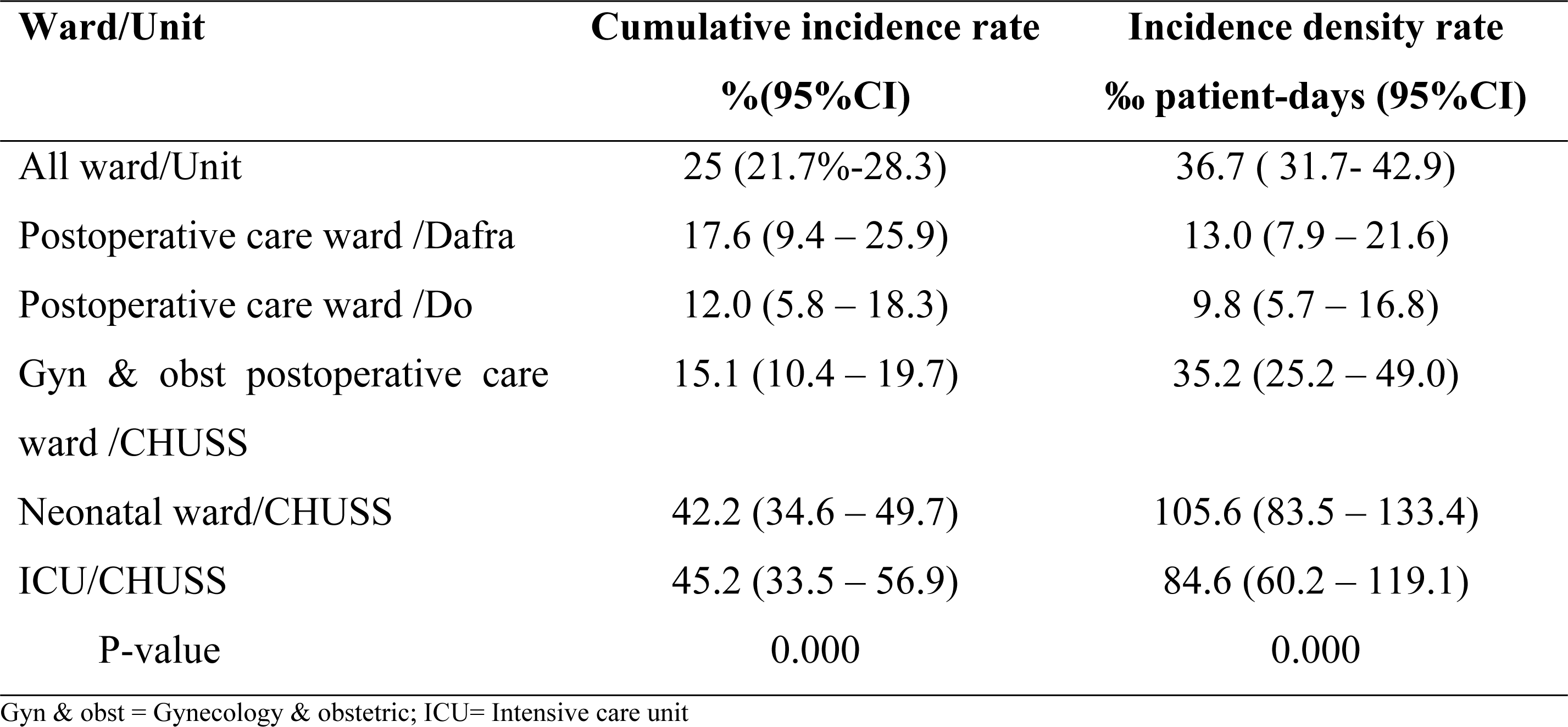
Incidence of healthcare-associated infection by ward specialty, multicenter longitudinal survey, Bobo-Dioulasso, 2022.

#### Typology of HAIs

Of the 166 HAIs, surgical site infections (SSIs) were the most common, accounting for 44.6% (74/166) of all HAIs and occurring in 16.0% of patients who underwent surgery (74/464). After SSIs, the next most common infections were neonatal infections, which accounted for 42.2% (70/166) of HAIs (Figure 1).

**Figure1:**
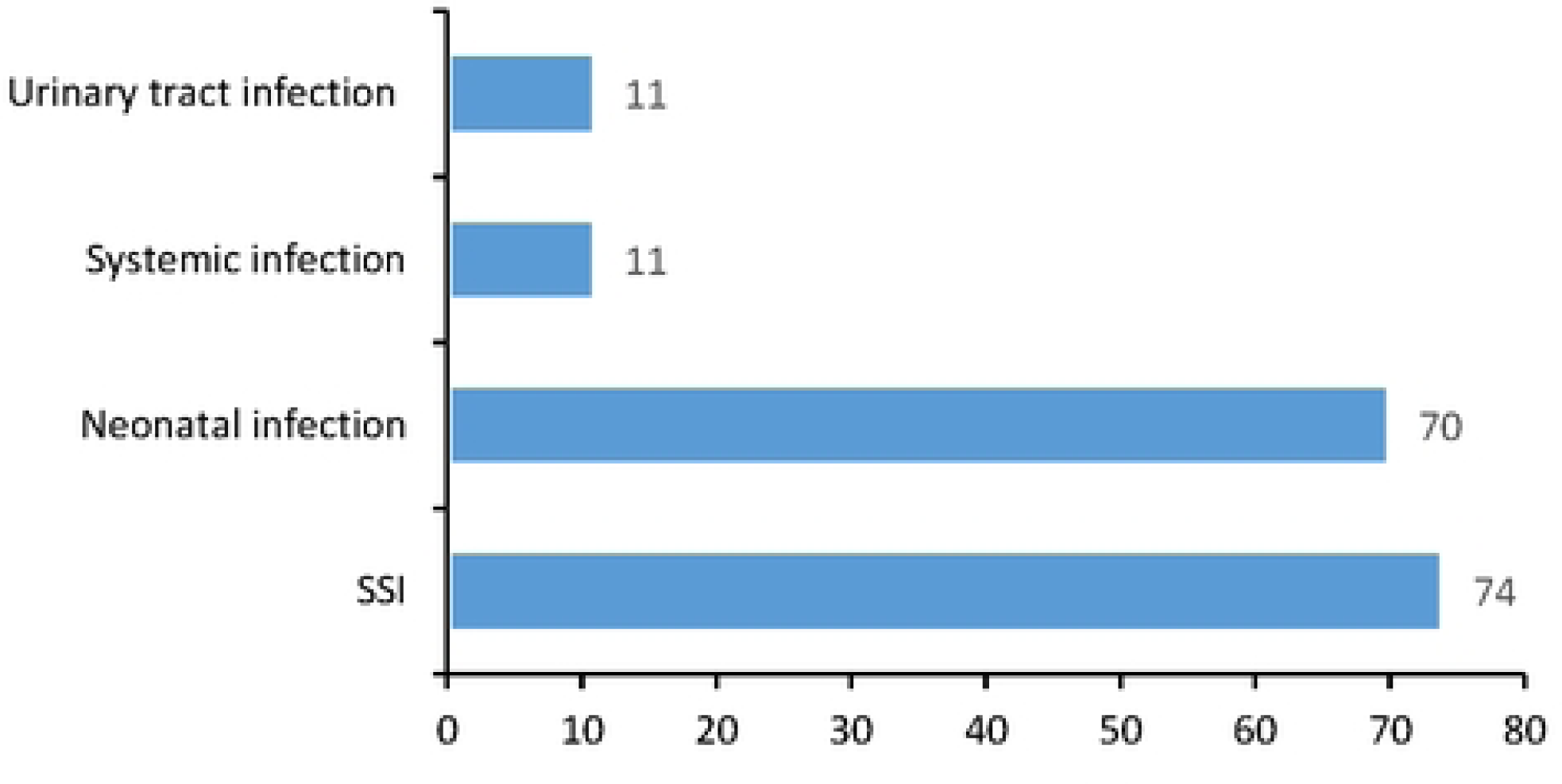
Distribution of healthcare-associated infection(n=166), multicenter longitudinal survey, Bobo-Dioulasso 2022.

### 3.3. Bacteriological profile of healthcare-associated infections

A total of 47 microorganisms were identified in 79 samples taken from 166 HAIs. The most common microorganisms found among the 45 were Escherichia coli (31.9%), *Klebsiella pneumoniae* (17%), *Acinetobacter baumannii* (12.8%), *Pseudomonas aeruginosa* (8.5%), *Staphylococcus epidermidis* (8.5%), *Staphylococcus aureus* (6.4%), *Enterococcus sp* (4.3%), *Klebsiella oxytoca* (4.3%), *Pantoea spp1* (2.1%). Figure 2 summarizes the distribution of different groups of microorganisms. Resistances to 3^rd^ generation cephalosporins, amoxicillin/clavulanate and amoxicillin were 51.8% (14/27), 69.2% (18/26) and 100% (13/13), respectively. The frequencies of resistance to cephalosporins and amoxicillin/clavulanate were highest in the neonatal ward (100% & 100%) and the ICU (66.7% & 100%). The proportion of extended-spectrum beta-lactamase producing Enterobacteriaceae (ESBLE) was 38.9% (7/28). They were isolated in the neonatal ward (n=2), in the ICU (n=3) and in the postoperative ward of Dafra (n=2). Of three (03) *Staphylococcus aureus* identified, one (01) was methicillin resistant (ICU) 33.3%.

**Figure 2:**
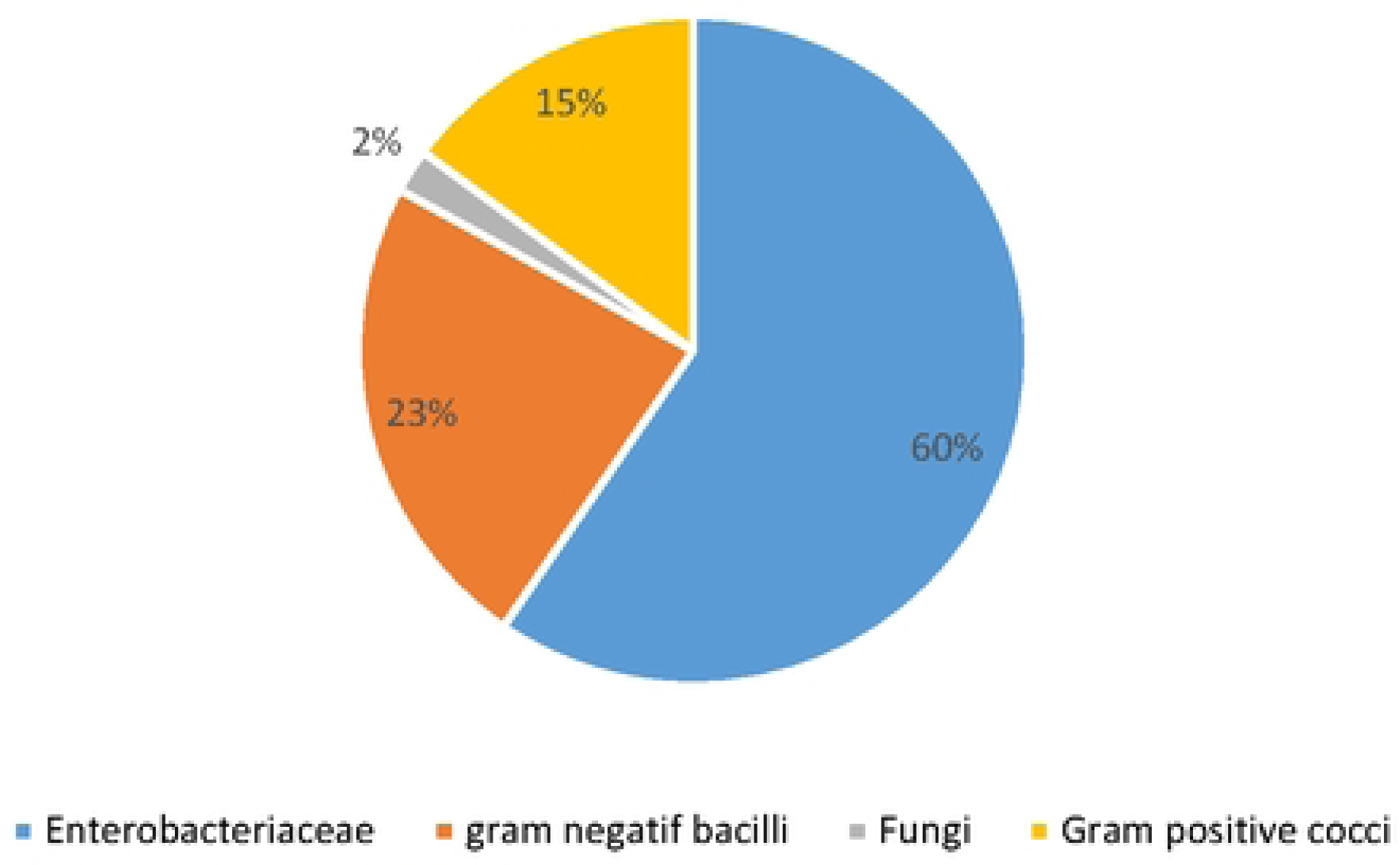
Groups of microorganisms (n=47), multicenter longitudinal survey, Bobo-Dioulasso 2022.

After multivariate analysis, only hospital stay of more than 2 days before enrollment (aOR=2.0; CI: 1.2-3.3), male sex (aOR=1.8; CI: 1.1-3.0), admission in neonatal ward (aOR=6.6; CI: 1.1-40.2) or ICU (aOR=3.3; CI: 1.3-8.5) were factors associated with a higher risk of HAI. There was no association between age group and HAIs (Table 3).

**Table 3:**
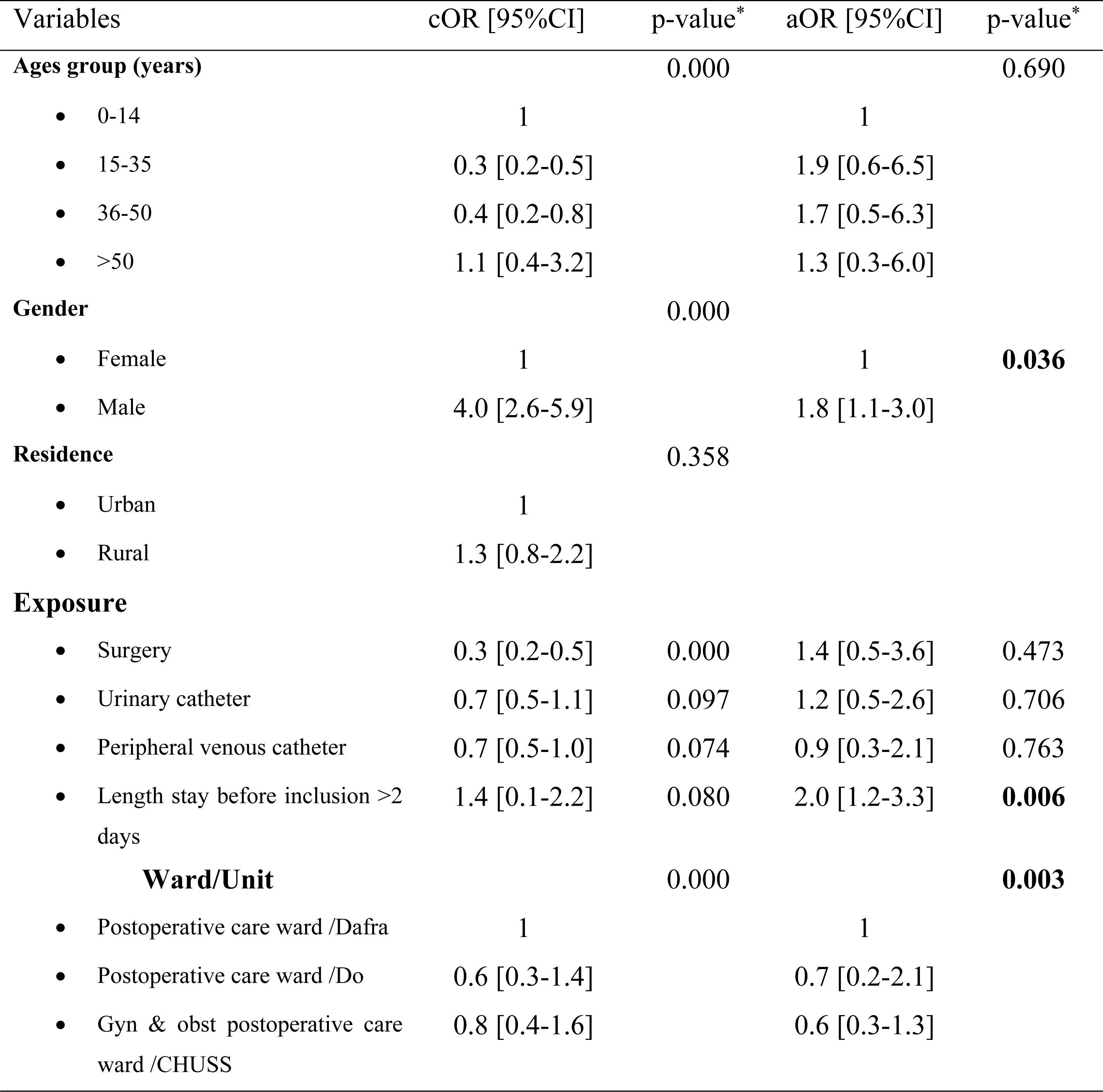
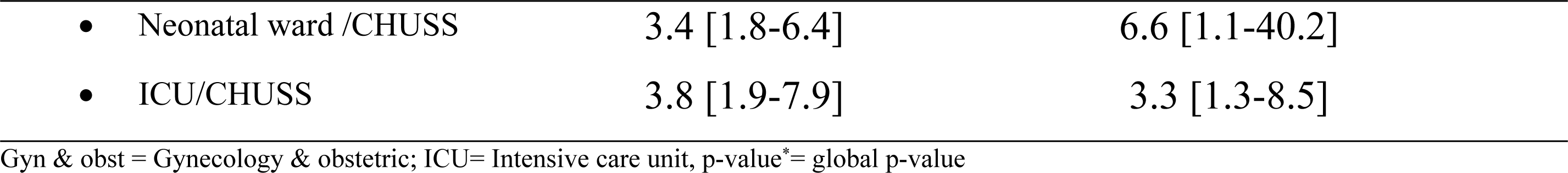
Healthcare-associated infection risk factor, multicenter longitudinal survey, Bobo-Dioulasso, 2022.

### 3.5. HAI Outcomes

The mean follow-up duration of included patients was 9.4 days (SD: 9.2) with a range of 2 to 30 days. It was twice as long for HAIs (15.7 days vs 7.3 days; p=0.000). The mean length of hospital stay was 7.8 days (SD: 7.9). It was longer for HAI cases (8.9 days vs 7.4 days; p=0.039). Of the 664 patients included, we observed 569 recoveries (85.7%), 6 discharges against medical advice (0.9%), 62 deaths (9.3%), and 27 lost to follow-up (4.1%). Deaths were observed only in the neonatal ward (n=42) and ICU (n=20). The incidence of death was higher in patients with HAI (18.1%; CI:12.1 - 24.4) than in patients without HAI (6.4%; CI: 4.3 - 8.5) p = 0.000. HAI mortality was 45.4% (CI: 27.5 - 63.4) in the ICU and 21.4% HAI mortality (CI: 11.6 - 31.3) in the neonatal ward (p=0.019).

## Discussion

Our study reported a cumulative incidence rate of HAIs of 25% (CI: 21.7%-28.3%) or an incidence density rate of 36.7 per 1000 patient-days (CI: 31.7-4.3). The highest incidences of HAIs were observed in the ICU (45.2%; CI: 33.5-56.9%) and in the neonatal ward (42.2%; CI: 34.6-49.7%). We observed a low efficiency of bacteriological investigations (47/79). Enterobacteriaceae accounted for 60% of the bacteria identified in HAIs, and 38.9% of these were extended-spectrum beta-lactamase (ESBL) producers. Factors associated with HAIs were hospital stay longer than 2 days (aOR=2.0; CI:1.2-3.3), male sex (aOR=1.8; CI:1.1-3.0), admission to the neonatal ward (aOR=6.6; CI:1.1-40.2) or ICU (aOR=3.3; CI:1.3-8.5). HAIs were associated with increased length of follow-up, hospitalization, and mortality (18.1%; 95%CI:12.1 - 24.4). HAI mortality was higher in the ICU (45.4%; CI: 27.5 - 63.4) than in the neonatal ward (21.4%; CI: 11.6 - 31.3) p=0.019.

The data from this study showing a high incidence of HAIs in the three main hospitals in Bobo Dioulasso are similar to those reported in previous studies in the African literature [12,18–21]. The frequency of HAIs obtained in our study remains higher than those reported in studies from both developed countries [2,22,23] and other African studies [10,11,16,24,25]. Consistent with our studies, the highest incidences of HAIs were reported in ICU and neonatal ward [12,18]. The frequencies of healthcare-associated infections vary greatly from one country to another. Then, in the same country from one hospital to another and vary from one department to another in the same hospital depending on the context. These frequencies may differ from one study to another depending on the case definitions and methodology used. It is recognized that longitudinal or incidence studies such as the present one offer greater precision in assessing the extent of these infections [14]. However, these studies are time-consuming and require more resources than the prevalence studies which are more commonly conducted. The lack of infection control policies and trained personnel and the overcrowding of our hospitals may explain the high incidence of HAIs in our study.

Microbiological tests were available for only 79 of the 166 infected patients (47.6%), and of these, at least one pathogen was identified in 47 (59.5%). This rate is consistent with that reported in the literature (41-86%) [26], but remains much higher than that reported by Ezzi et al. in Tunisia [27]. Systematic empirical antibiotic therapy, whether prophylactic or curative, sometimes prior to bacterial culture, a common practice in the neonatal ward, could explain the poor performance of culture in our study. However, the predominance of enterobacteria in HAIs has been widely reported in the literature [5,6,19,28,29] and was confirmed in our study. In addition, this study highlighted a relatively high prevalence of EBLSE 38.9%, which is consistent with data from the literature [5,6,28]. These findings suggest the need to rationalize the use of antibiotics in our hospitals by establishing therapeutic protocols based on antimicrobial resistance surveillance data. The risk factors for HAI in our study are consistent with the results of numerous studies showing an association between length of hospital stay, male sex, neonatal or ICU admission and the occurrence of these infections [10,18,30]. Long hospital stays mean prolonged exposure to pathogens in the hospital environment, as well as an increased likelihood of invasive medical procedures, resulting in a higher risk of HAI. The association of male sex with an increased risk of HAIs is a controversial topic. However, differences between men and women in terms of personal hygiene practices, of comorbidities frequency and anatomy may explain this association [32,33,34]. Patients admitted to neonatal and ICU are often vulnerable due to their compromised immune system. In addition, the high number of critically ill patients and the complex medical procedures performed in these units make them a breeding ground for pathogens.

Our study, like many others, shows an association between HAIs and increased length of hospital stay, comorbidity and high mortality [2]. Our results show an overall HAI mortality of 18.1% (95%CI: 12.1 - 24.4), which is lower than the 28.7% reported by Latifa Merzougui et al. in Tunisia [19]. Our case fatality rate of 45.4% (95% CI:27.5 - 63.4) in ICU HAIs is similar to the case fatality rate of Latifa Merzougui et al (44.7%) [19] and that of Shahnaz Rimaz in the Islamic Republic of Iran (45.2%) [29]. Data on HAI outcomes vary according to study methodology, accuracy of HAI case identification strategy, the typology of HAIs studied, and the quality of care. Prospective, longitudinal studies such as ours are more appropriate than retrospective studies for assessing HAI outcomes.

The main limitation of this study was its relatively short inclusion period (2 to 4 months), which does not allow analysis of seasonal variations in the incidence of HAIs. In addition, the present study does not take into account environmental and behavioral factors of healthcare workers in the analysis of risk factors of HAIs. However, its prospective longitudinal design has the advantage of allowing the analysis of the etiological factors of HAIs.

### Conclusion

The incidence of HAIs was relatively high in the three hospitals in Bobo Dioulasso. The pathogens identified in these infections were mainly Enterobacteria, with more than one-third being EBLS. Risk factors for HAI included prolonged stay, male sex, neonatal or ICU admission. This study underlines the need to implement a policy for the prevention and control of these infections, focusing on the training of healthcare personnel, surveillance of HAIs and the development of therapeutic protocols for the rational use of antibiotics.

## DECLARATIONS

### Ethics approval and consent to participate

The study was approved by the National Health Ethics Committee of Burkina Faso. An information sheet was given to each participant. After reading this information sheet, those who desired to participate in the study signed a free and informed consent form. The data collected were anonymized to ensure their confidentiality.

### Data availability

The datasets, including deidentified provider quantitative and qualitative data, used and/or analyzed during the current study are available from the corresponding author upon reasonable request.

## Acknowledgments

We would like to thank the data collectors, the staff of Sourô Sanou Teaching Hospital, Do and Dafra District Hospitals. Our gratitude extends to the patients for their participation in this study.

## Supporting information

**S1_Figure1:** Distribution of healthcare-associated infection(n=166), multicenter longitudinal survey, Bobo-Dioulasso 2022

**S2_Figure 2:** Groups of microorganisms (n=47), multicenter longitudinal survey, Bobo-Dioulasso 2022

**S1_Table 1:** Characteristics of patients, multicenter longitudinal survey, Bobo-Dioulasso 2022

**S2_Table 2:** Incidence of healthcare-associated infection by ward specialty, multicenter longitudinal survey, Bobo-Dioulasso, 2022

**S3_Table 3:** Healthcare-associated infection risk factor, multicenter longitudinal survey, Bobo-Dioulasso, 2022

